# High Accuracy Machine Learning Model for Sarcopenia Severity Diagnosis based on Sit-to-stand Motion Measured by Two Micro Motion Sensors

**DOI:** 10.1101/2023.05.18.23289933

**Authors:** Keer Wang, Hongyu Zhang, Clio Yuen Man Cheng, Meng Chen, King Wai Chiu Lai, Calvin Kalun Or, Yonghua Chen, Yong Hu, Arul Lenus Roy Vellaisamy, Cindy Lo Kuen Lam, Ning Xi, Vivian W. Q. Lou, Wen Jung Li

## Abstract

In this ageing society, sarcopenia as a geriatric condition that can have significant negative impacts on an individual’s quality of life. Sarcopenia is a kind of aged syndrome associated with loss of muscle mass and function, which may lead to falls, fractures, gait disorders or even mortality. There are multiple ways to diagnose sarcopenia, such as using Magnetic resonance imaging (MRI), Dual-energy X-ray absorptiometry (DEXA) and Bioelectrical impedance analysis (BIA) etc. to calculate muscle mass; using handgrip or sit-to-stand to measure muscle strength; using short physical performance battery (SPPB), gait, and 5-time sit-to-stand to evaluate physical performance.

In this work, we use two μIMUs worn on subjects to record their sit-to-stand motion, and then used several machine learning models to diagnose the severity of sarcopenia of the subjects. We recruited 53 elderly subjects in total for this work. The youngest subject is 65 years old and the oldest is 84 years old. Their average age is 70 years old. Among these 53 subjects, there are 12 healthy ones and 41 sarcopenia patients with different severity. The subject is instructed to do the single sit-to-stand (STS) three times, and two μIMUs attached to the subject’s waist and thigh transfer the data to a computer by Bluetooth. We separated the STS motion process into 4 phases based on the angle and angular velocity, extracted a total of 510 features for motion analytics. These features were futher analyzed by sequential feature selection with 5 different machine learning models (SVM, KNN, decision tree, LDA, and multilayer perceptron). With our proposed methodology, all 53 subjects could be classified as healthy or having sarcopenia with risk level 1, 2, or 3. The best accuracy to distinguish the healthy or sarcopenia subjects is 98.32%, and the best results to distinguish sarcopenia risk levels from 0 (healthy) to 3 (most severe) is 90.44%.

## I. Introduction

Sarcopenia is a progressive and generalized loss of skeletal muscle mass and strength syndrome due to ageing [1]-[3]. Both muscle mass and muscle strength will begin to decrease after a person reaching a certain age. Typically, muscle mass decreases at a rate of 3-5% per decade after 30 years old, and the speed of decrease accelerates a lot after age 60 [4]. Risks of adverse outcomes like frailty, fall, physical disability, and mortality also increase due to sarcopenia. The prevalence of sarcopenia rises with ageing and differs across different settings and clinical conditions. Diagnosing sarcopenia early and getting treatments as early as possible is important.

Different research groups have different definitions of sarcopenia [5]-[11]. There are three main consideration components of the sarcopenia diagnosis in summary: muscle mass, muscle strength and physical performance.

To measure muscle mass, there are mainly six methods: Anthropometry, Bioelectrical impedance analysis (BIA), Dual-energy X-ray absorptiometry (DEXA), Computer tomography (CT), Magnetic resonance imaging (MRI) and Ultrasound [1], [10]-[15]. Anthropometry is a simple and low-cost technique easily applied in clinical practice and large population-based surveys. Yet, the sensitivity of changes may be limited due to different observers. BIA is a method for estimating body composition, in particular, body fat and muscle mass. It is safe and has no radiation exposure [17]-[19]. However, it has a significant disadvantage in that the muscle mass measurements can be distorted by hydration status and the presence of oedema. DEXA is usually used as a means of measuring bone mineral density (BMD), but also sometimes used to assess body composition and fat content [20][21]. Even though the radiation exposure is small, it is expensive, time-consuming and has poor accessibility. CT scan is a medical imaging technique that can obtain detailed internal body images [22]. It can review the images after scanning. Since CT scan mainly utilizes X-ray, it is radiation exposure and requires interpretation by a radiologist. It also needs a confined space for the scanner. MRI is similar to CT but uses strong magnetic fields, magnetic field gradients and radio waves to generate organ images. It cannot use if the patient has metalwork/some pacemakers [23]. Ultrasound is a real-time medical imaging technique to visualize muscles, tendons and many internal organs [24][25]. It has no ionizing radiation and is highly safe. However, there are no criteria for diagnosis of low muscle mass with ultrasound.

To measure muscle strength, researchers mainly use handgrip strength, knee flexion/extension and peak expiratory flow [7]. Handgrip strength is a simple and convenient measure of muscle strength, and it is widely used in clinical practice. It strongly correlates with lower extremity muscle power, knee extension torque and calf cross-sectional muscle area [26]-[28]. To measure the knee flexion/extension, the subjects are instructed to sit in an adjustable straight-back chair, the lower leg unsupported and the knee flexed to 90 degrees and the force applied to the ankle will be recorded [29]. The knee flexible/extension is suitable for research but not very convenient in clinical practice for the need for special equipment and training. Peak expiratory flow measures the strength of respiratory muscles, but it is not recommended as an isolated measure of muscle strength for sarcopenia diagnosis [7].

To measure physical performance, Short Physical Performance Battery (SPPB), Gait, and Sit-to-stand (STS) are usually instructed [30]-[32]. SPPB is an objective assessment tool for evaluating lower extremity function in the elderly [33]. It involves a series of physical performance tests, including balance tests (stand side by side, semi-tandem and tandem), gait, and STS [33][34]. It is a standard physical performance measurement for both research and clinical practice. Both the gait and STS tests are part of the SPPB, but they can also be used individually as a single parameter for clinical practice and research. The STS test varies with different research, such as single sit-to-stand (STS), the 5-time sit-to-stand (5TSTS), 30s chair stand and timed get-up-and-go [35]-[39].

A micro inertial measurement unit (μIMU) is an electronic sensing device that can be used to measure the body’s specific acceleration, angular rate, and orientation [41]. Because the μIMUs are sensitive, low cost and portable [44]-[46], they are attracting increased scientific attention for motion analysis on disease diagnosis. Cuzzolin et al. utilized an IMU to diagnose Parkinson’s based on gait analysis and achieved an accuracy of 85.51% over Parkinson’s patients and healthy subjects [47]. Pham and his group mainly detect Parkinson’s through the sit-to-stand and stand-to-sit transitions with one IMU attached to the lower back [48]. Witchel et al. also assess the sit-to-stand and stand-to-sit transition with IMUs but in multiple sclerosis to measure the patient’s postural control [49]. Ko et al. in 2021 evaluate the physical performance of the elderly by time-up-and-go test and gait to diagnose sarcopenia [50]. Kim’s group analyzed the gait data with 2 IMUs on feet and can receive an accuracy of 88.69% to identify osteopenia and 93.75% for sarcopenia [51]. This work uses a μIMU to record sit-to-stand motion of subjects and applied machine learning related algorithms to different patients with different sarcopenia severity.

## II. Severities of sarcopenia

There are many different groups to diagnose sarcopenia, but most of them just diagnose whether the subject has sarcopenia or not. However, some organizations also proposed different classifications of the severity of sarcopenia. EWGSOP introduced three conceptual stages of sarcopenia: pre-sarcopenia (low muscle mass), sarcopenia (low muscle mass and low muscle strength/low physical performance) and severe sarcopenia (low muscle mass, low muscle strength and low physical performance) [7]. EWGSOP2 in 2018 updated the operational definition of sarcopenia: probable sarcopenia (low muscle strength), sarcopenia (low muscle strength and low muscle quantity or quality), and severe sarcopenia (low muscle strength, low muscle quantity or quality and low physical performance) [5]. AWGS 2019 extended the diagnosis method from AWGS 2014 following extensive deliberations. They defined possible sarcopenia by low muscle strength with or without reduced physical performance; sarcopenia by low muscle mass, low muscle strength or low physical performance; severe sarcopenia by low muscle mass, low muscle strength and low physical performance.

The inclusion criteria are from the diagnosis approach of AWGS 2019. The cutoffs from the AWGS 2019 are shown below. Low muscle strength is defined as handgrip strength <28 kg for men and <18 kg for women; criteria for low physical performance are 6-m walk <1.0 m/s, Short Physical Performance Battery score ≤9, or 5-time chair stand test ≥12 seconds. The cutoffs for height-adjusted muscle mass are dual-energy X-ray absorptiometry, <7.0 kg/m2 in men and <5.4 kg/m2 in women, and bioimpedance, <7.0 kg/m2 in men and <5.7 kg/m2 in women [40].

The AWGS 2019 method is mainly used to diagnose whether the subject is healthy or not. A new definition of different levels of severities of sarcopenia is proposed based on AWGS 2019 method. There are three tasks from the physical performance part in the AWGS 2019 method: 6-metre walk, 5-time chair stand test and short physical performance battery (SPPB). Each task has a criterion: 6-metre walk < 1.0 m/s; 5-time chair stand test ≥ 12s; SPPB ≤ 9. Each criterion is a risk. If the subject meets one of these three, he/she is defined as risk level 1; If the subject meets any two of these three, he/she is defined as risk level 2; If the subject meets all these three, he/she is defined as risk level 3.

## III. Experiment Design

To record the subjects’ motion sit-to-stand, we utilize two μIMU sensors. The μIMU sensors we selected are commercial devices from WitMotion Co. Ltd (China). Its product model is BWT901CL. This kind of μIMU is small size, low-cost, portable, wireless and easy to operate. The IMU’s dimension is 51 × 36 mm and 15 mm thick. The weight of the μIMU is only 20 grams. The parameters of the μIMU are shown in Table 1. We utilize the two 9-axis μIMUs to record the movement of sit-to-stand. The full-scale ranges of the accelerometer, gyroscope and magnetometer are ± 16g, ± 2000 °/*s* and ± 2 Gauss, respectively. The sampling rate is 200 Hz. This kind of μIMU is chargeable and once it is fully charged, it can last for 4 hours. The data can be transferred to the computer by Bluetooth 2.0.

**Table 1.**
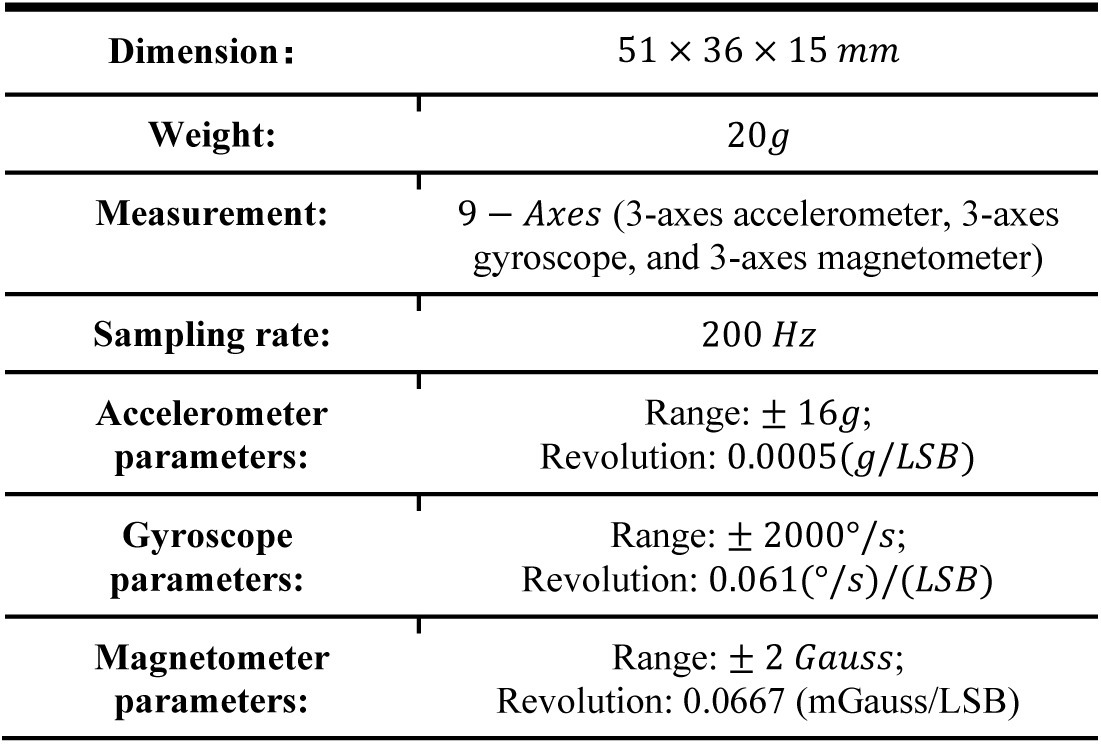
Basic parameters of the μIMU (BWT901CL)

|  |  |
| --- | --- |
| **Dimension:** | 51 × 36 × 15 mm |
| **Weight:** | 20g |
| **Measurement:** | 9 – Axes (3-axes accelerometer, 3-axes gyroscope, and 3-axes magnetometer) |
| **Sampling rate:** | 200 Hz |
| **Accelerometer parameters:** | Range: $\pm 16g$;<br>Revolution: 0.0005(g/LSB) |
| **Gyroscope parameters:** | Range: $\pm 2000^\circ/s$;<br>Revolution: 0.061($^\circ/s$)/(LSB) |
| **Magnetometer parameters:** | Range: $\pm 2$ Gauss;<br>Revolution: 0.0667 (mGauss/LSB) |

We recruited 53 elderly subjects in total for this work. The youngest subject is 65 years old and the oldest is 84 years old. The average age is 70 years old. Among these 53 subjects, there are 12 healthy ones and 41 sarcopenia patients with different severity. According to the risk level definition mentioned previously, there are 12 patients in risk level 1, 6 patients in risk level 2 and 23 patients in risk level 3, making up the total of 41 sarcopenia patients. The subject demographics are shown in Table 2.

**Table 2.**
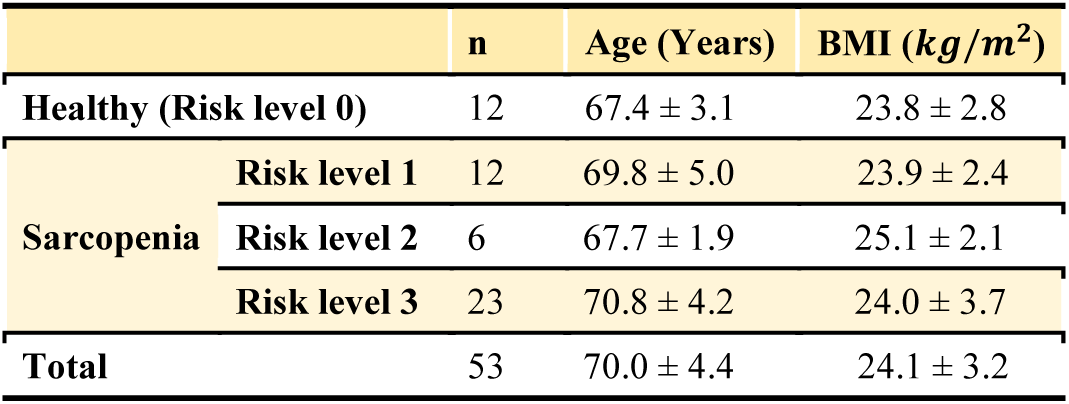
Summary of participant demographics.

The two μIMUs were attached to the subject’s waist and thigh with belts separately, as shown in Fig. 2. The subjects were instructed to sit on an armless chair crossing their arms in front of the chest and then stand up. The height of the chair is 45cm. The subjects were not allowed to lean back on the chair or to assume a perched position. If the subject could complete the motion without any help, they could finish the motion with armrests. Each subject was asked to conduct the sit-to-stand process three times.

**Fig. 1.**
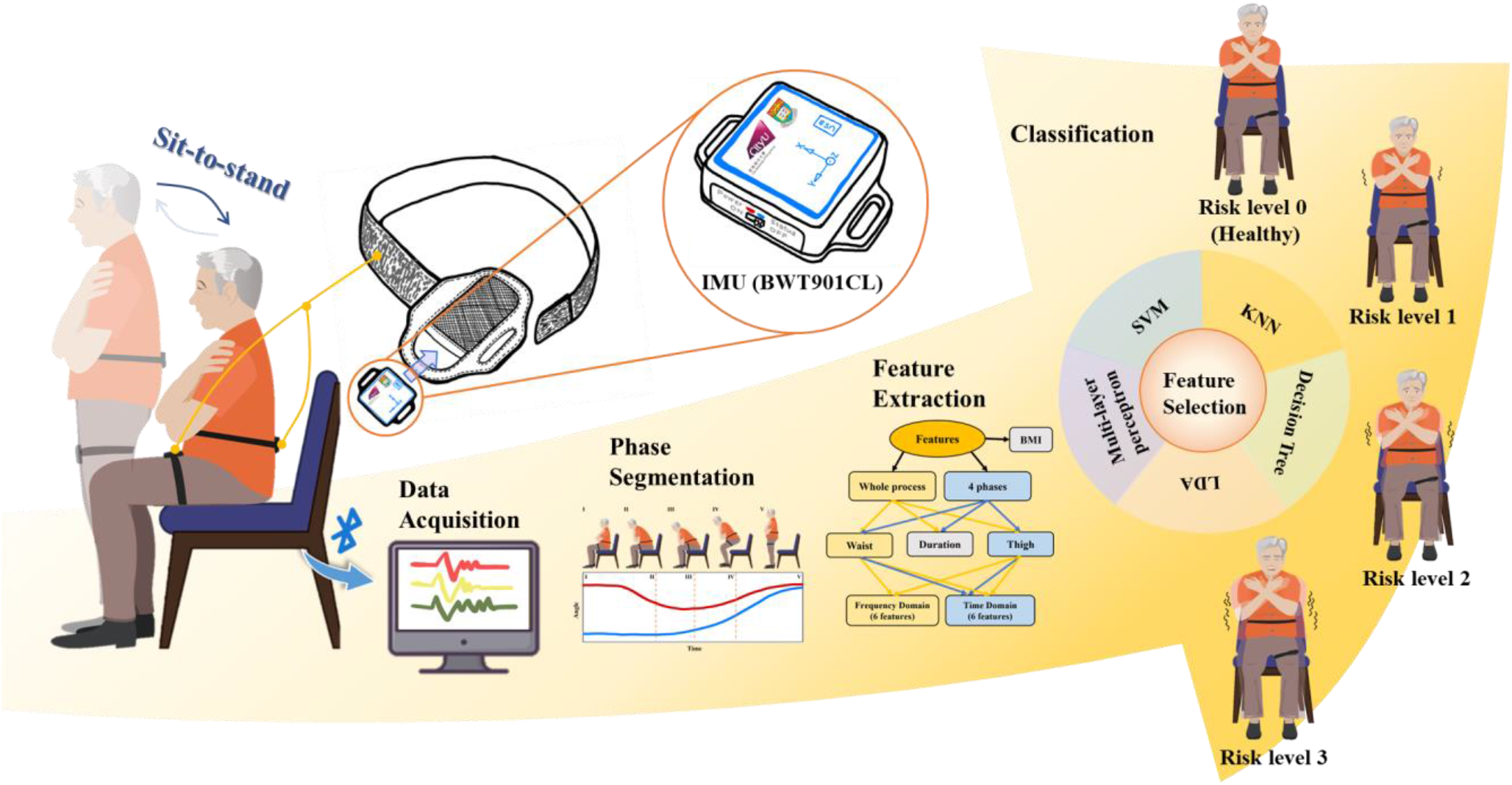
The experimental process of sarcopenia diagnosis in this work.

**Fig. 2.**
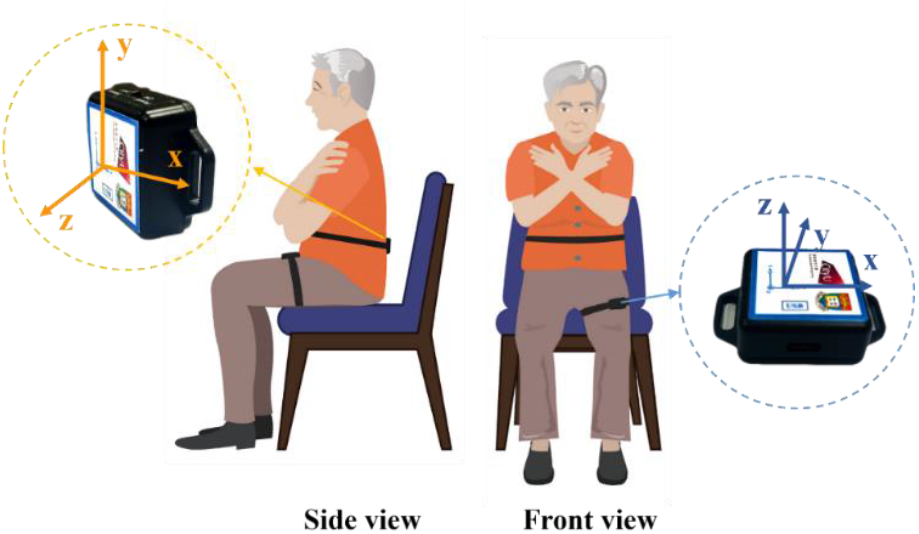
Waist senor orientation has the x-axis put horizontally and pointing to the subject’s left, the y-axis is vertical and points up, and the z-axis is in the sagittal plane with the direction parallel to the horizontal axis and points frontally. The thigh sensor’s orientation is the same as the waist sensor.

## IV. Data Analysis

### A. Preprocessing

Since there are two μIMUs here, it’s crucial to synchronize. The thigh data is recorded first and then the waist data. When the subject finishes the motion, the waist data is stopped first and then the waist data. The total data recorded from the thigh μIMU is more than that from the waist μIMU. In order to synchronize the two series of data, we cut the thigh data according to the absolute time.

To detect the starting moment, the sliding window is utilized. The length of the window is 40. OW is the standard deviation of the sliding window. “*OW* > (0.05 ∗ *the mean value of OW*)” can be viewed as a criterion. The first point to reach this criterion is defined as the starting point of sit-to-stand. The last point to reach this criterion is defined as the ending point of sit-to-stand. Fig. 3 shows the curve changes from 2 μIMUs during the single sit-to-stand.

**Fig. 3.**
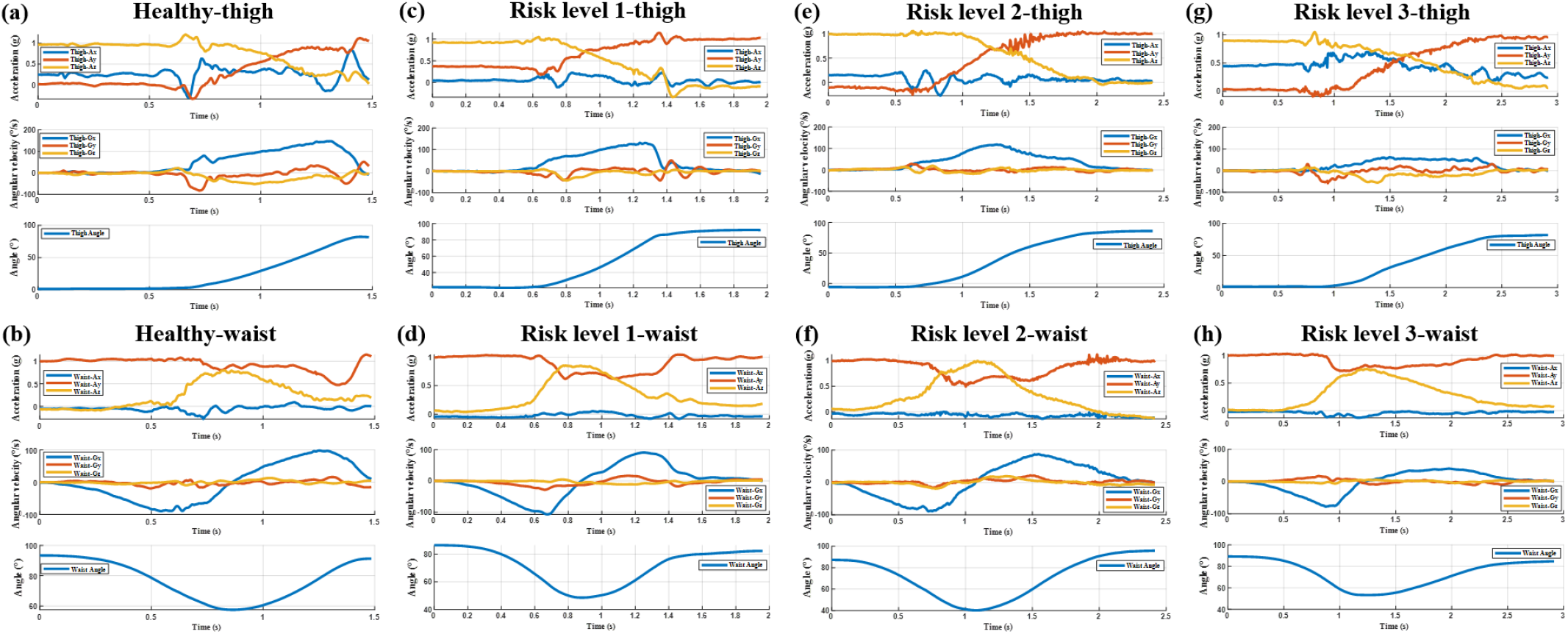
(a) The sit-to-stand data of a healthy subject from the μIMU positioned on waist; (b) The sit-to-stand data of a healthy subject from the μIMU positioned on the thigh; (c) The sit-to-stand data of the subject with Risk level 1 from the μIMU positioned on waist; (d) The sit-to-stand data of the subject with Risk level 1 from the μIMU positioned on the thigh; (e) The sit-to-stand data of the subject with Risk level 2 from the μIMU positioned on waist; (f) The sit-to-stand data of the subject with Risk level 2 from the μIMU positioned on the thigh; (g) The sit-to-stand data of the subject with Risk level 3 from the μIMU positioned on waist; (h) The sit-to-stand data of the subject with Risk level 3 from the μIMU positioned on the thigh.

### B. Phase Segmentation

Sit-to-stand is fundamental and essential in daily life. Many researchers have analyzed this motion and divided it into different phases. Among all these phase division methods, there are three fixed positions: Sitting, Seat-off and Standing. The Sitting position refers to the initial position of the motion of sit-to-stand. The Seat-off position refers to the position that the subject’s body leaves the chair (there is no extra force applied to the chair). The Standing position refers to the end position of the motion of sit-to-stand.

In this paper, we divide the sit-to-stand motion into 5 critical points (4 phases) based on the collected μIMU data, as shown in Fig. 4. The main reference of the division method is the angular velocity and angle curves. These five points are identified as

**Fig. 4.**
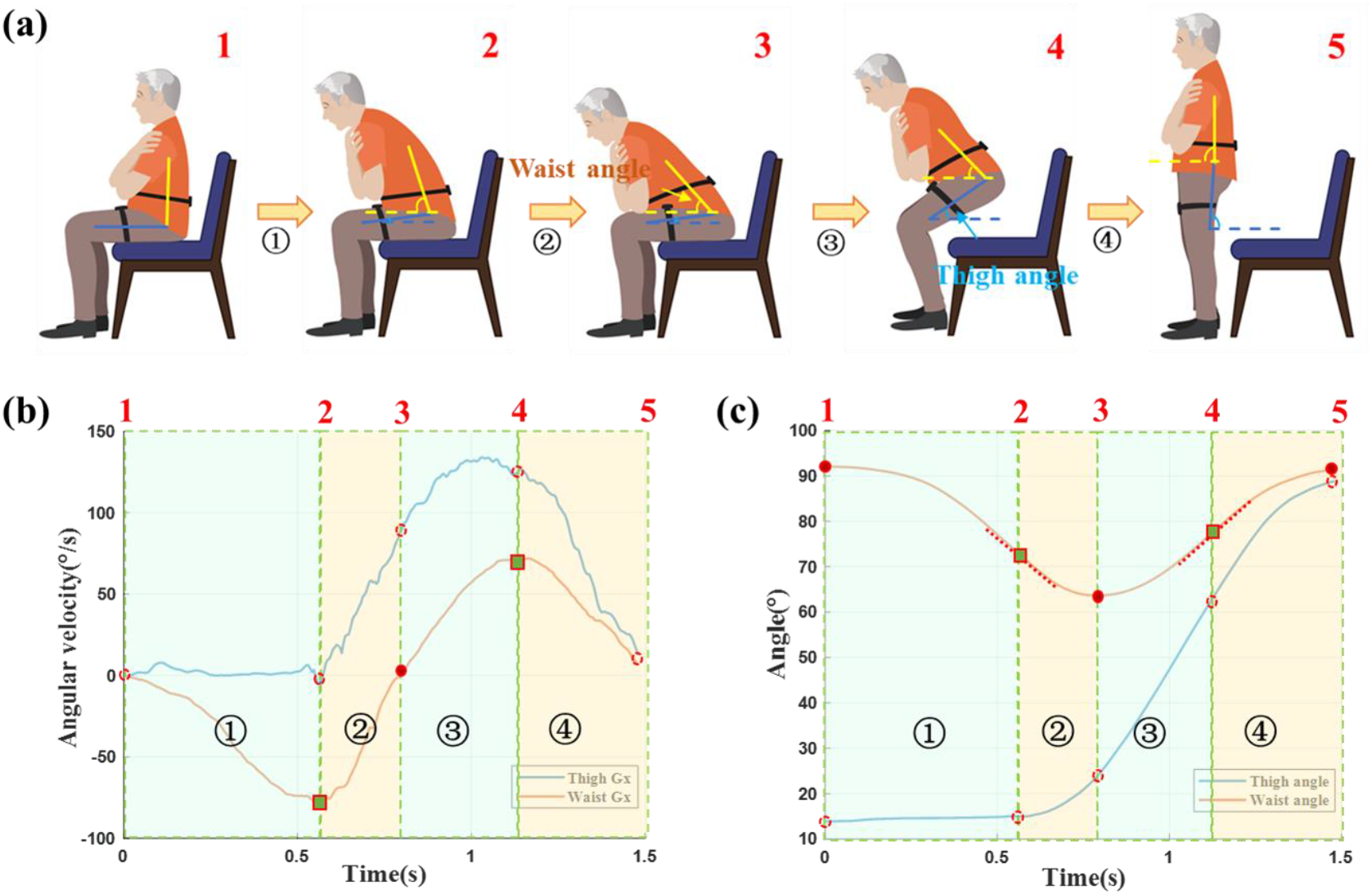
The sit-to-stand motion can be segmented into 5 critical points (4 phases). (a) Shows five segmentation points of sit-to-stand; (b) shows the corresponding points on the curves of the x-axis of angular velocity of waist and thigh; (c) shows the corresponding points on the curves of the waist angle and thigh angle.

1. STS initiation;
2. Peak flexion angular velocity;
3. Seat off;
4. Peak extension angular velocity;
5. STS termination.

STS initiation is defined by the starting point of sit-to-stand introduced in Preprocessing. Peak flexion angular velocity is the trough of the X-axis of angular velocity from the waist μIMU. Seat off is the point that the X-axis of angular velocity from the waist μIMU equals zero or the peak of the waist angle. Peak extension angular velocity is the crest of the X-axis of angular velocity from the waist μIMU. STS termination is defined by the ending point of sit-to-stand introduced in preprocessing.

### C. Feature Extraction

The features we extracted are from the sit-to-stand whole motion, 4 phases and personal information, as shown in Fig. 5. In the sit-to-stand part, we extract features from both the frequency domain and time domain. In the 4 phases part, we extract features from the time domain. Both the sit-to-stand part and 4 phases part include the features from the 7-axis of 2 μIMUs and their corresponding durations. For personal information, we calculate the BMI of each subject and take it as a feature.

**Fig. 5.**
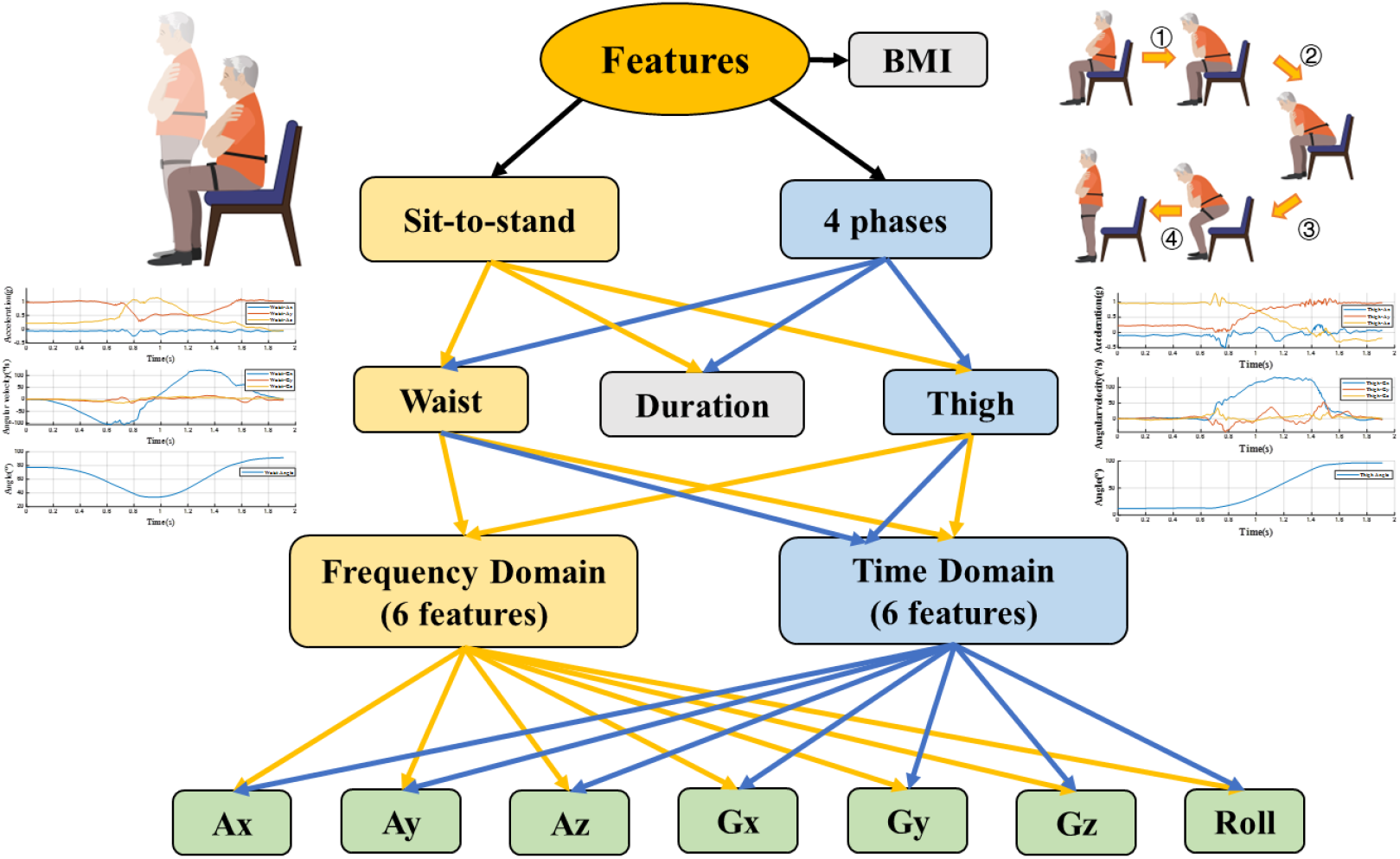
Feature extraction from the STS and 4 phases.

The features extracted from the time domain and frequency domain are shown in Table 3. The features in the time domain include max, min, mean, peak to peak, standard deviation and root mean square. The features in the frequency domain include average frequency, Barycenter frequency, mean square frequency, frequency root mean square, frequency variance and frequency standard deviation.

**Table 3.** Statistical and morphological features.

| Domain | Feature | Formula |
| --- | --- | --- |
| Time Domain | Max | $x_{max} = \max_{n=1,2,...,N} \{x_n\}$ |
| | Min | $x_{min} = \min_{n=1,2,...,N} \{x_n\}$ |
| | Mean | $\bar{x} = \frac{1}{N} \sum_{n=1}^N x_n$ |
| | Peak to peak | $x_{p-p} = x_{max} - x_{min}$ |
| | Standard deviation | $\sigma_x = \sqrt{\frac{1}{N-1} \sum_{n=1}^N [x_n - \bar{x}]^2}$ |
| | Root mean square | $x_{rms} = \sqrt{\frac{1}{N} \sum_{n=1}^N x_n^2}$ |
| Frequency Domain | Average frequency | $FA = \frac{1}{K} \sum_{k=1}^K S_k$ |
| | Barycenter frequency | $FC = \frac{\sum_{k=1}^K f_k S_k}{\sum_{k=1}^K S_k}$ |
| | Mean square frequency | $MSF = \frac{\sum_{k=1}^K f_k^2 S_k}{\sum_{k=1}^K S_k}$ |
| | Frequency root mean square | $RMSF = \sqrt{MSF}$ |
| | Frequency variance | $VF = \frac{\sum_{k=1}^K (f_k - FC)^2 S_k}{\sum_{k=1}^K S_k}$ |
| | Frequency standard deviation | $RVF = \sqrt{VF}$ |

We separate these features into two feature sets: STS and STS+4 phases. The features of STS are 170 in total (2 μIMU × (6 Time Domain + 6 Frequency Domain) × 7 Axes + BMI + Sit-to-stand Duration = 170). The features of STS + 4 phases are 510 in total ([2 μIMU × (6 Time Domain + 6 Frequency Domain) × 7 Axes + S it-to-stand Duration] + [2 μIMU × 4 Phases × 6 Time Domain + 4 × Phase Duration] + BMI = 510).

### D. Machine Learning

#### 1) Support Vector Machines (SVM)

SVM is a supervised two-class classifier. It is a general feed-forward neural network of traditional machine learning. The basic model of SVM is to find the best-separating hyperplane in the feature space to maximize the interval between positive and negative samples on the training set.

The interval of the function represents the certainty that the feature is positive or negative, given the training samples (*x*^(*i*)^, *y*^(*i*)^), then we can get

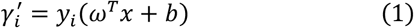

The geometric interval is the distance from the vector point to the overclocking surface. The equation is as following:

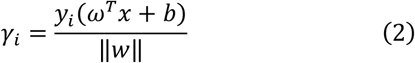

Suppose there is a set of training samples with two classifications (+1 and -1), the decision surface is

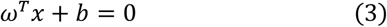

The *x* is the input vector, *w* is the adjustable weight vector, and *b* is the bias.

Suppose the model is linear, then

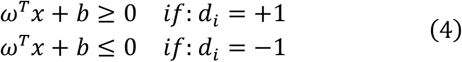

Margin is twice the interval *γ* from the hyperplane to the nearest data point. If a plane can maximize *γ*, it is called the optimal hyperplane.

Suppose the weight and bias have optimal solution, the optimal hyperplane function and the corresponding discriminant function are

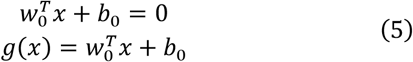

Then the (*w*_0_, *b*_0_) in the sample set *x*^(*i*)^ must satisfy:

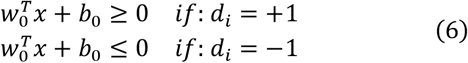

The (*x*_*i*_, *d*_*i*_) is the support vector. These points are closest to the hyperplane. The distance between the points *x* and the optimal hyperplane is

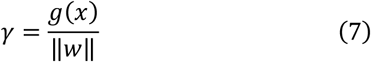

In the positive and negative hyperplanes, any support vectors satisfying the following equation:

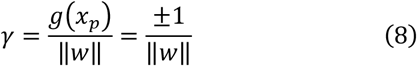

Margin can be defined as:

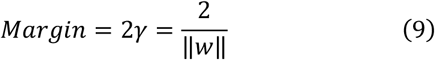

In order to maximize *γ*, we need to minimize ∥*w*∥ . Maximize the margin between two class equals to minimize the Euclidean norm of weight vector *w*.

#### 2) K-nearest Neighbor (KNN)

KNN is a machine learning technique and algorithm that can be used for both regression and classification tasks. KNN examines the labels of a chosen number of data points surrounding a target data point, in order to make a prediction about the class that the data point falls into. A KNN model calculates similarity using the distance between two points on a graph. The greater the distance between the points, the less similar they are. A KNN algorithm can be illustrate as following:

a. Setting K to the chosen number of neighbors.
b. Calculating the distance between a provided/test example and the dataset examples.
c. Sorting the calculated distances.
d. Getting the labels of the top K entries.
e. Returning a prediction about the test example.

In the first step, K is chosen individually and it tells the algorithm how many neighbors (how many surrounding data points) should be considered when rendering a judgment about the group the target example belongs to. In the second step, the model checks the distance between the target example and every example in the dataset. The distances are then added into a list and sorted. Afterward, the sorted list is checked and the labels for the top K elements are returned. When rendering a prediction about the target data point, it matters if the task is a regression or classification task. For a regression task, the mean of the top K labels is used, while the mode of the top K labels is used in the case of classification.

#### 3) Decision Tree (DT)

A decision tree is a decision that goes from the root node to the leaf node step by step. It can be applied to both classification and regression. The leaf node is the final decision result.

The metric for feature segmentation is entropy. Entropy is the measure of how disorder the random variables is. Entropy is proportional to uncertainty. The equation of entropy is

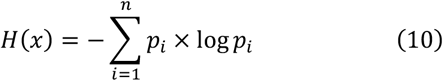

Where *p*_*i*_ is the probability of case *i*. When the probability is 0 or 1, the random variable has an uncertainty relation of 1. When the probability is 0.5, the random variable is max.

The information gain indicates how the feature reduces the uncertainty relationship of the class, and the information gain index is usually used as the selection criterion of the node decision feature.

#### 4) Linear Discriminate Analysis (LDA)

LDA is a supervised machine learning that project the data into a low-dimensional space, making the same type of data as compact as possible. The algorithm of LDA is as following.

Considering a set of data *D* = (*x*_1_, *y*_1_), (*x*_2_, *y*_2_), …, (*x*_*m*_, *y*_*m*_), *x*_*i*_ is a n-dimensional vector, class *y*_*i*_ belongs to *C*_1_, *C*_2_, …, *C*_*k*_. We define the *N*_*j*∈1,2,…,*k*_ the number of sample class j, *X*_*j*(*j*∈1,2,…,*k*)_ the collection of sample class j, *μ*_*j*(*j*∈1,2,…,*k*)_ the average of sample class j.

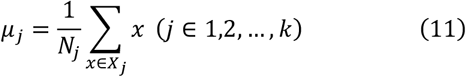

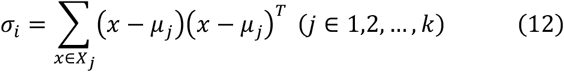

The between-class scatter matrix is

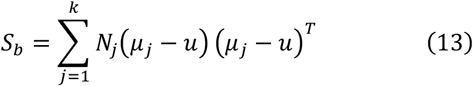

Where u is the average value of all the data.

The Intra-class scatter matrix is

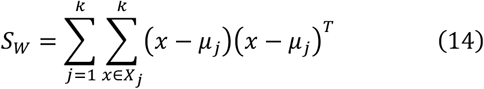

We project the low-dimensional space dimension into d-dimension. The corresponding basis vector is *ω*_1_, *ω*_2_, …, *ω*_*d*_,, forming a matrix W. Function J can be defined as below:

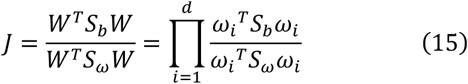

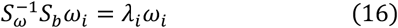

When equation (31) is existed, the function has maximum value. We take the eigenvectors corresponding to the first *d* largest eigenvalues to form matrix *W*. Since *S*_*b*_ is the addition of *k* matrices with a rank of 1, its rank is less than or equal to *k*. And because we know that after the first (*k* − 1) of *μ*_*j*_, the last *μ*_*k*_ can be represented by the first (*k* − 1). The highest dimension after the dimensionality reduction of the LDA algorithm is (*k* − 1).

#### 5) Multilayer perceptron (MLP)

A multilayer perceptron (MLP) is a fully connected class of feedforward artificial neural network (ANN). An MLP can be viewed as a directed graph consisting of multiple layers of nodes, each fully connected to the next layer. Except for the input node, each node is a neuron (or processing unit) with a nonlinear activation function. MLPs are usually trained by a supervised learning method known as the backpropagation algorithm. It generally consists of at least three layers of nodes: an input layer, a hidden layer and an output layer.

The algorithm for the MLP can be described as follows:

a. As with the perceptron, the inputs are passed through the MLP by taking the dot product of the input with the weights between the input layer and the hidden layer (WH). This dot product gives a value in the hidden layer.
b. MLPs make use of activation functions at each of their computed layers. There are many activation functions to discuss: rectified linear units (ReLU), sigmoid function, and tanh. Pass the computed output at the current layer through one of these activation functions.
c. Once the calculated output at the hidden layer has been pushed through the activation function, push it to the next layer in the MLP by taking the dot product with the corresponding weights.
d. Repeat step b and c until the output layer is reached.
e. At the output layer, the calculations are either used for a backpropagation algorithm according to the activation function selected for the MLP (in the case of training), or a decision is made based on the output (in the case of testing).

MLPs form the basis of all neural networks and have greatly increased the power of computers when applied to classification and regression problems. Thanks to the multilayer perceptron, computers are no longer limited by XOR cases and can learn rich and complex models.

### E. Feature Selection

Sequential feature selection (SFS) can be used to identify the most problem-relevant features from all features. With SFS, we can reduce an initial a-dimensional feature space to a b-dimensional feature subspace (b<a) by using the greedy search algorithm. Fig. 6 shows the flow chart of SFS. We use 5 different machine learning models (SVM, KNN, DT, LDA, MLP) to select the 510 features. The selection criterion is the accuracy of the classification. The steps of SFS are as follows:

**Fig. 6.**
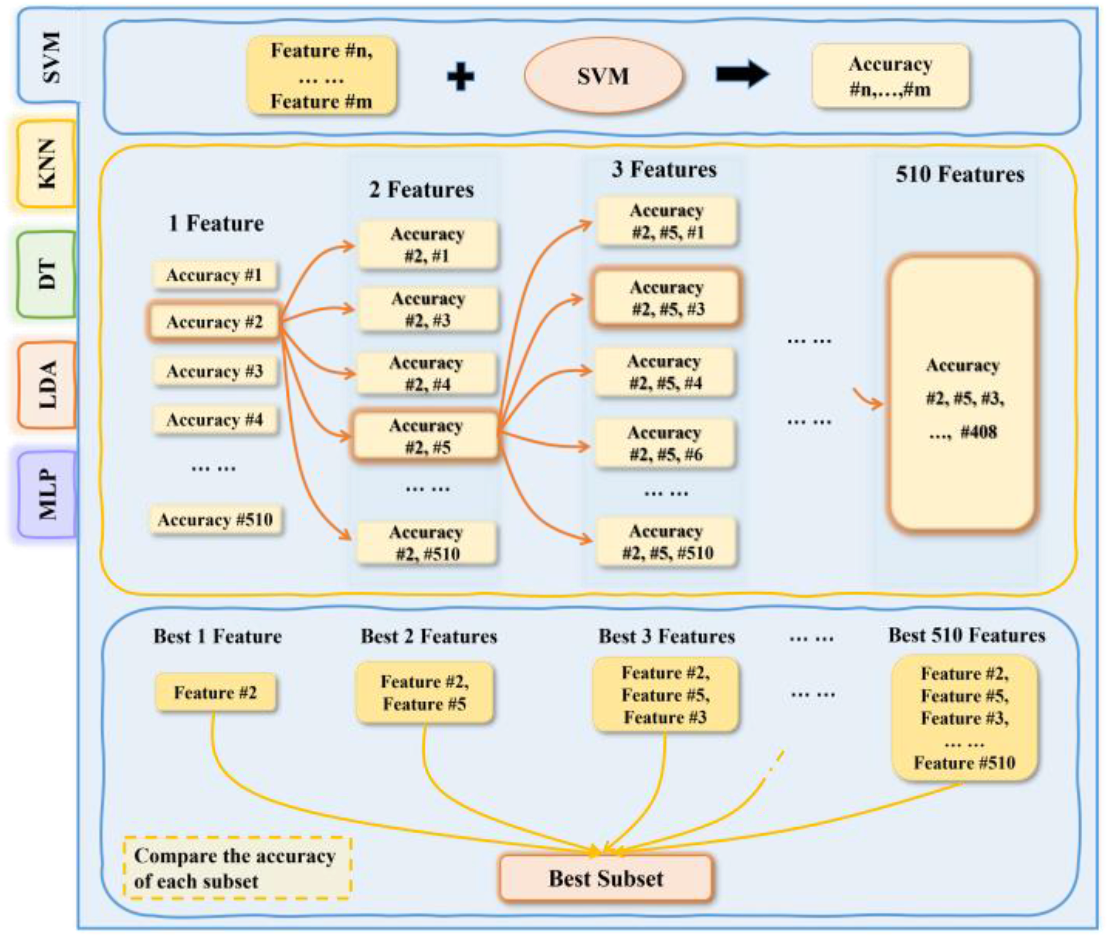
Sequential Feature Selection (SFS).

a. At the first time, one feature is put in the machine learning model, and it comes out as the best 1 feature for the classification.
b. Hold this feature, and group it with the other 509 features respectively as the input of the second time machine learning.
c. Repeat the steps until the input is 510 features.
d. Compare the accuracies of the 510 groups of features, the group with the highest accuracy is chosen as the best subset.

We define the two classifications of healthy and sarcopenia subjects as Group 1, and the 4 classifications of risk level 0-3 subjects as Group 2. Table 4 shows the results of feature selection by SFS. We can see that in the comparison of Group 1, the DT method selects the fewest features (8 features), and the LDA method selects the most features (269 features). The SVM, KNN and MLP methods select 20 features, 11 features and 28 features respectively. To classify risk level 0-3, the method of DT, SVM and MLP choose 11 features, 20 features and 36 features. The features selected by the LDA and KNN methods for the comparison of Group 2 are a bit more, 65 features and 84 features respectively.

**Table 4.**
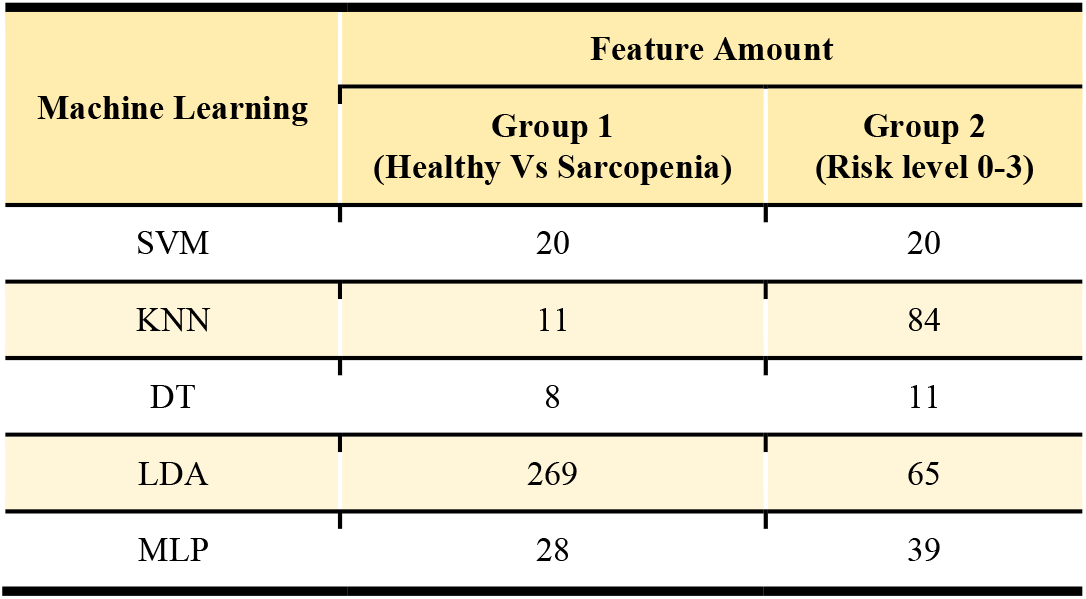
Results of feature selection

| Machine Learning | Feature Amount |  |
| --- | --- | --- |
|  | Group 1<br>(Healthy Vs Sarcopenia) | Group 2<br>(Risk level 0-3) |
| SVM | 20 | 20 |
| KNN | 11 | 84 |
| DT | 8 | 11 |
| LDA | 269 | 65 |
| MLP | 28 | 39 |

## IV. Results

Sarcopenia is a kind of disease that is quite prevalent for the elderly population beyond 60 years of age. The decreases in muscle mass and muscle strength easily lead to falls and bone fractures which are quite dangerous to the elderly. So, it is essential to diagnose sarcopenia and its severities early. In this paper, we utilize 2 μIMUs to analyze the movement of sit-to-stand for diagnosing different severity of sarcopenia.

We proposed a novel sit-to-stand phase segmentation method based on μIMU data. By dividing the process of sit-to-stand, we could extract more features and better analyze the motion.

We put the 2 feature sets (STS and STS+4 phases) into five classification models (SVM, LDA, DT, KNN and MLP) separately for different group classifications. The feature set STS has 170 features in total mainly from the motion of sit-to-stand. The feature set of STS+4 phases has 510 features in total from both the motion of sit-to-stand and 4 phases of the motion. For the classification models, the rate of the training set and testing set is 7:3 and the cross-validation is 10 folds.

The results shown are the average accuracies of 5 times classifications.

Fig. 7 shows the results of feature extraction. In the classification of healthy and sarcopenia subjects (Group 1), feature set STS can get the highest accuracy of 93.34% by KNN among five classification models; The feature set STS+4 phases can get the highest accuracy of 96.66% by MLP among five classification models. In the classification of risk level 0-3 subjects (Group 2), both the STS and STS+4 phases feature sets get the highest accuracy 90.44% and 85.82% respectively by using the classification model KNN.

**Fig. 7.**
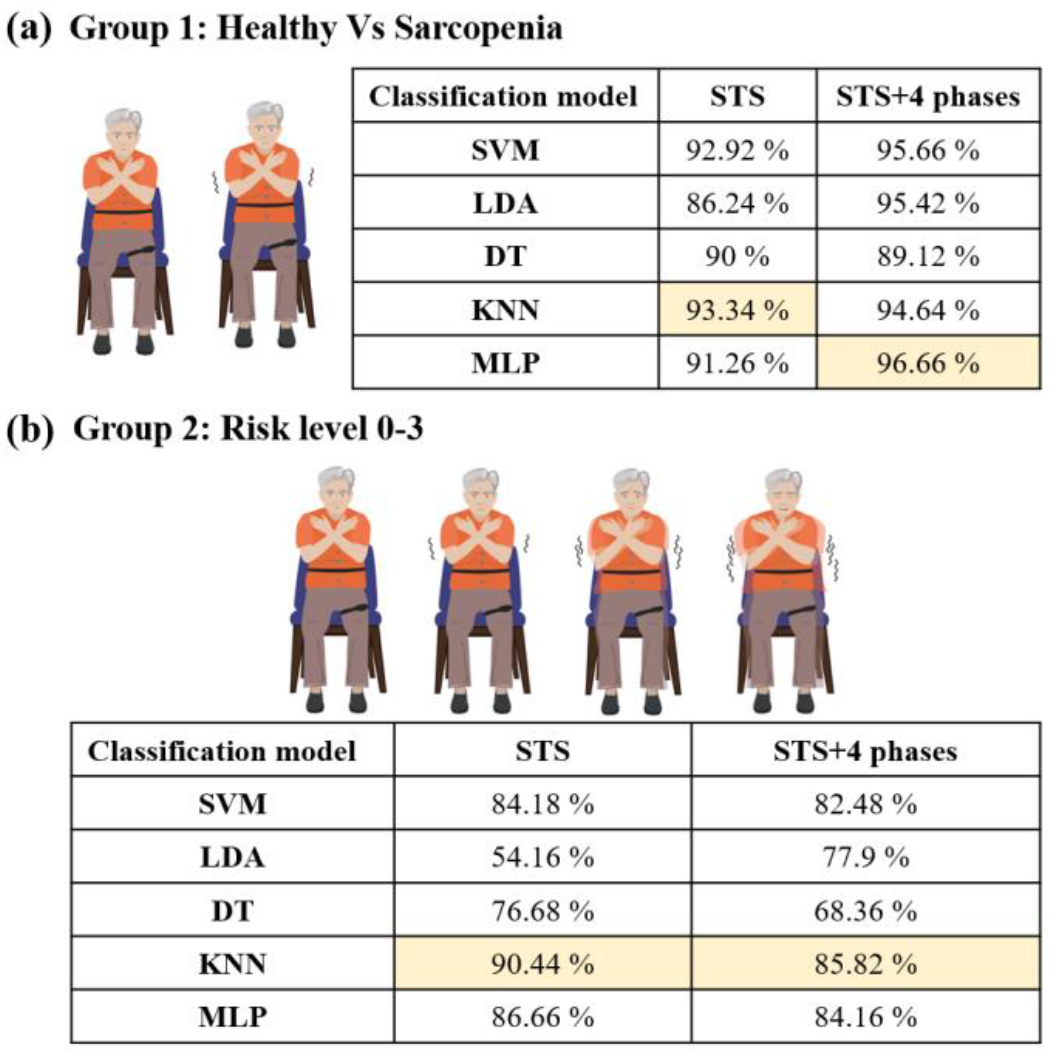
The results of the feature extraction. (a) shows the accuracies of STS and STS+4 phases with 5 classification models (SVM, LDA, DT. KNN and MLP) in the comparison of healthy and sarcopenia subjects. (b) shows the accuracies of STS and STS+4 phases with 5 classification models in the comparison of risk level 0-3 subjects.

We select features from 510 features by sequential feature selection. Fig. 8 shows the results of feature selection and classifications. The first blue boxes in Fig. 7 (a) and (b) show the classification results with features selected out by SFS with SVM. The yellow boxes show the classification results with features selected out by SFS with KNN. The green boxes show the classification results with features selected out by SFS with DT. The red boxes show the classification results with features selected out by SFS with LDA. The purple boxes show the classification results with features selected out by SFS with MLP. In each box, there are five bars which refer to the accuracies obtained from the five classification models (SVM, KNN, DT, LDA and MLP).

**Fig. 8.**
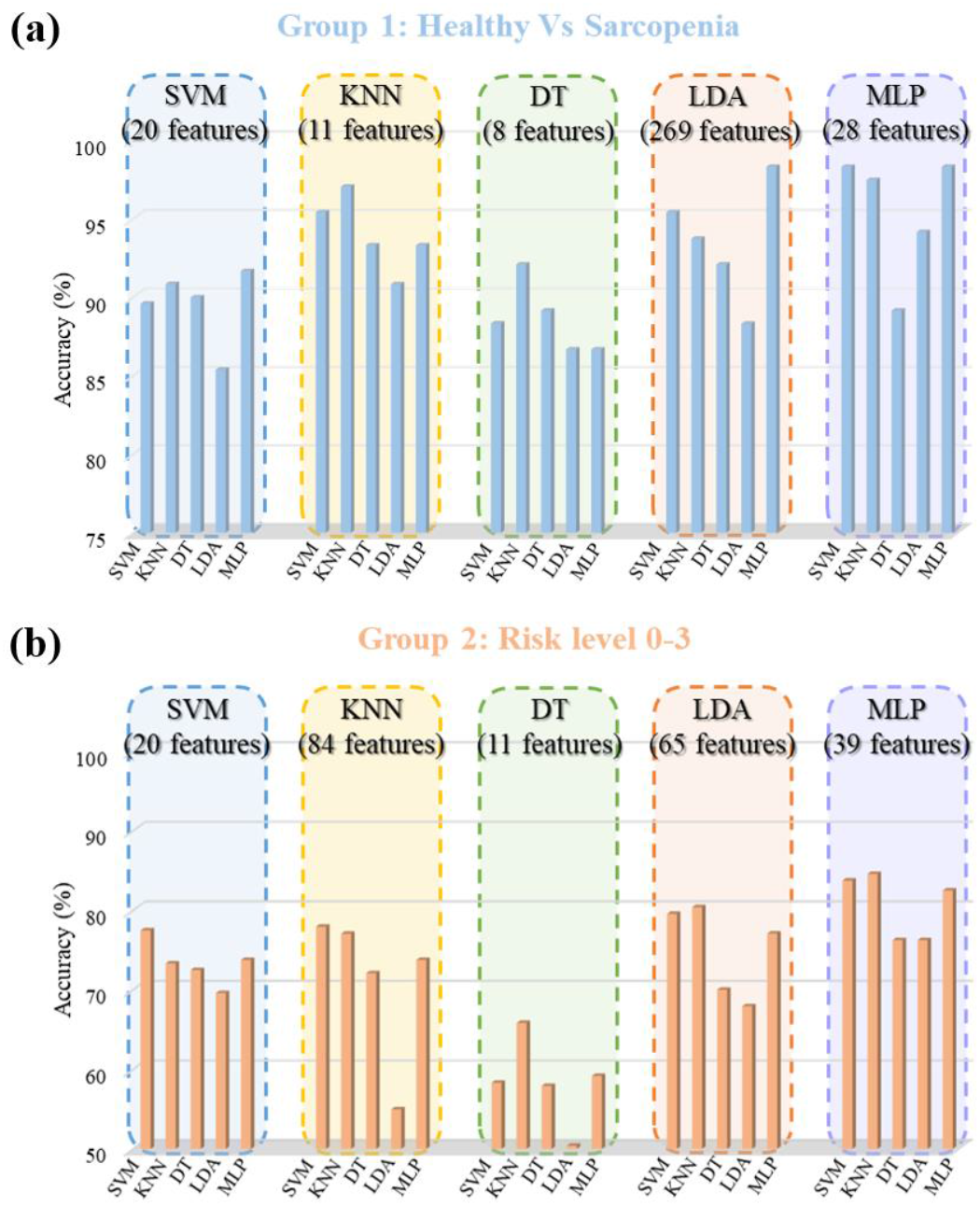
The results of feature selection. (a) Shows the accuracies calculated by 5 feature sets selected out by feature selection with 5 classification models (SVM, LDA, DT. KNN and MLP) in the comparison of healthy and sarcopenia subjects. (b) shows the accuracies calculated by 5 feature sets selected out by feature selection with 5 classification models in the comparison of risk level 0-3 subjects.

In Group 1, to classify the healthy and sarcopenia subjects, we can see that feature set selected out by MLP (28 features) leads to the highest accuracy 98.32% using the classification SVM model. In Group 2, to classify the risk level 0-3 subjects, the feature set selected out by MLP (39 features) leads to the highest accuracy 84.58% calculated by the classification model KNN.

In summary, the highest accuracy obtained in classifying the healthy and sarcopenia subjects is 98.32%, which was obtained by using the SVM classification model with 28 features selected out by SFS; the highest accuracy obtained in classifying the risk level 0-3 subjects is 90.44% using the KNN classification model feature set STS (170 features).

## VI. Conclusion

In this work, we presented our results on diagnosing the severity of sarcopenia of 41 subjects by sit-to-stand motion with two wearable μIMUs. We proposed a novel way to analyze the motion of sit-to-stand using 2 μIMUs attached on the subjects’ thigh and waist and separating the motion into 4 phases. In total, 510 features can be extracted from the entire sit-to-stand motion and the 4 phases. With the help of machine learning models, we can classify the healthy and sarcopenia subjects with a high accuracy of 98.32%. Moreover, we have obtained an accuracy of 90.44% in distinguishing the sarcopenia subjects of three different risk levels 1, 2, and 3.

## Data Availability

All data produced in the present study are available upon reasonable request to the authors.

## Data Availability

All data produced in the present study are available upon reasonable request to the authors.

